# Risk for hypertension and heart failure linked to high normal serum sodium and tonicity in electronic medical records

**DOI:** 10.1101/2024.07.05.24309996

**Authors:** Jonathan Rabinowitz, Mahmoud Darawshi, Nuriel Burak, Manfred Boehm, Natalia I Dmitrieva

## Abstract

**Background and Aims:** Population aging is fueling an epidemic of age-related chronic diseases. Managing risk factors and lifestyle interventions have proven effective in disease prevention. Epidemiological studies have linked markers of poor hydration with higher risk of chronic diseases and premature mortality. Many individuals do not adhere to recommended hydration levels and could benefit from improved hydration habits. Our study evaluates the use of electronic medical records to confirm the relationship between inadequate hydration and the risk of chronic diseases, which may inform hydration-focused interventions in general healthcare.

**Methods:** We analyzed 20-year electronic medical records for 411,029 adults from Israel’s Leumit Healthcare Services. Hydration status was assessed using serum sodium and tonicity. We included adults without significant chronic diseases or water balance issues, defined as having normal serum sodium (135-146 mmol/l) and no diagnosis of diabetes. We used Cox proportional hazards models, adjusted for age, to assess the risk of developing hypertension and heart failure.

**Results:** Our findings showed an increased risk of hypertension with elevated serum sodium levels: a 12% rise for the 140-142 mmol/l group and 30% for levels above 143 mmol/l (HR1.30, 95%CI:1.26-1.34). Tonicity over 287 mosmol/kg was associated with a 19% increased risk of hypertension (HR1.19, 95%CI:1.17-1.22). The risk of heart failure also increased, reaching 20% for sodium levels above 143 mmol/l (HR1.20,95%CI:1.12-1.29) and 16% for tonicity above 289 mosmol/kg (HR1.16, 95%CI: 1.10-1.22). The association between sodium and hypertension was observed across genders, while the risk of heart failure was more pronounced in females. Within the healthy Leumit cohort, 19% had serum sodium levels within the 143-146 mmol/l range, and 39% were in the 140-142 mmol/l range.

**Conclusions:** Data analysis from electronic medical records identified a link between serum sodium of 140 mmol/l and above and increased risk of hypertension and heart failure in the general Israeli population. Identifying individuals with high-normal sodium values in healthcare records could guide improvements in hydration habits, potentially leading to better health outcomes.

## Introduction

Population aging is driving a global epidemic of age-dependent chronic diseases, including cardiovascular, chronic respiratory, musculoskeletal, neurological, and mental disorders. It is estimated that these disorders in individuals aged 60 years and older account for approximately 23% of the total global disease burden, and this proportion is rapidly increasing as life expectancy rises.^1^ Consequently, identifying mechanisms and implementing preventive measures to decelerate the aging process have emerged as new challenges for biomedical research and public health ^1^. In response to these challenges, geroscience and lifestyle medicine have arisen as rapidly growing disciplines that study the role of lifestyle factors in the prevention and treatment of chronic diseases^2,3^.

Identifying and addressing risk factors before the onset of diseases or early in the course of disease progression has proven effective in delaying or preventing the development of diseases. For instance, the recognition of hypertension as a significant risk factor for most chronic diseases, coupled with its widely adopted treatment, is considered a major public health success that has substantially reduced the prevalence of and mortality from cardiovascular and other diseases^4,5^. Additionally, strong evidence from intervention trials supports the conclusion that primary preventive lifestyle measures—such as a healthy diet, regular physical activity, stress management, and smoking cessation—offer preventive benefits that are often superior to those of pharmacotherapy (reviewed in ^2^).

Recent findings suggest that optimal hydration could serve as a preventive strategy to decelerate aging and reduce the risk of chronic diseases. This hypothesis originates from several longitudinal epidemiological studies that have associated indicators of poor hydration—such as elevated plasma copeptin levels, increased serum sodium, tonicity, and osmolality, as well as reduced urine volume and higher urine osmolality—with a heightened risk of chronic diseases and early death (reviewed in ^6^). Support for this hypothesis comes from an intervention study in mice; lifelong water restriction shortened their lifespan and led to degenerative changes that suggest accelerated aging ^7^. Although human interventional trials are necessary to substantiate these findings (with recent trials underway^8^, reviewed in ^6^), immediate action on hydration is justifiable. Global surveys indicate that approximately half of the population does not meet the existing general guidelines for optimal hydration ^9^.

Identifying risk factors through rigorous epidemiological studies is just the first step. The ultimate goal is to mitigate these risks with population-level interventions. This involves establishing criteria to pinpoint increased risks, crafting recommendations, identifying at-risk individuals within populations, and encouraging adherence to these guidelines.

In this study, we leveraged electronic medical records from Leumit Healthcare Services, one of Israel’s four healthcare providers. Our aim was to determine if associations between hydration indicators and chronic disease risk, similar to those reported in controlled epidemiological studies, could be observed in routine medical practice data. We focused on serum sodium as our primary hydration marker, given its widespread testing in general medical practice, making it a globally accessible indicator for identifying at-risk individuals. We analyzed the risk of heart failure in relation to serum sodium, similar to the analysis previously performed in the Atherosclerosis Risk in Communities (ARIC) study ^10^. Additionally, we explored the link between serum sodium and the risk of hypertension, a strong risk factor for heart failure and most chronic diseases, which has much earlier onset and widely accessible diagnostic procedures. Our analysis serves as a model for similar studies worldwide, using medical records to investigate hydration as a risk factor and to implement hydration-focused interventions within primary care systems..

## Methods

### Population and dataset

The dataset analyzed in the current study was provided by Leumit Healthcare Services (hereafter Leumit), one of four national health funds under universal health care in Israel. Leumit provides medical coverage to 7.5% of the total population of Israel (about 720,000 members, 9.2 million patient visits a year) country wide. They have accumulated over 20 years of electronically documented medical records covering all patient visits. In this dataset, sample selection bias is minimized, because it is illegal for healthcare service organizations in Israel to refuse membership based on demographic factors, health conditions, or medication needs. Ethical approval to get access to the deidentified dataset and conduct the current study was granted by the Leumit Helsinki Institutional Review Board (protocol LEU-0009-23).

### Inclusion and exclusion criteria

Dataset that we requested and was provided by Leumit included males and females above the age of 18 years, who had at least two serum sodium alongside fasting blood glucose measurements after age of 18 years. We excluded people who had diagnosis of major chronic diseases at the time of first sodium test or were diagnosed within following 2 years: Heart failure (ICD9 - 428.x), Dementia (ICD-9 codes - 290.x), Chronic Obstructive Pulmonary Disease (ICD-9 codes -491.x), Asthma (ICD-9 codes - 493.x), Chronic Pulmonary Heart Disease (ICD-9 codes - 416.x), Cardiac Dysrhythmias ((ICD-9 codes - 427.x), Peripheral Vascular Disease ((ICD-9 codes - 443.x), Diabetes Mellitus (ICD-9 codes - 250.x), Stroke(ICD-9 codes - 436.3), Renal Failure (ICD-9 codes - 585.x; 586). Persons with a diagnosis of HIV at any time were also excluded. The final dataset provided by Leumit after these exclusions, contained 483,390 individuals (Figure 1).

**Figure 1.**
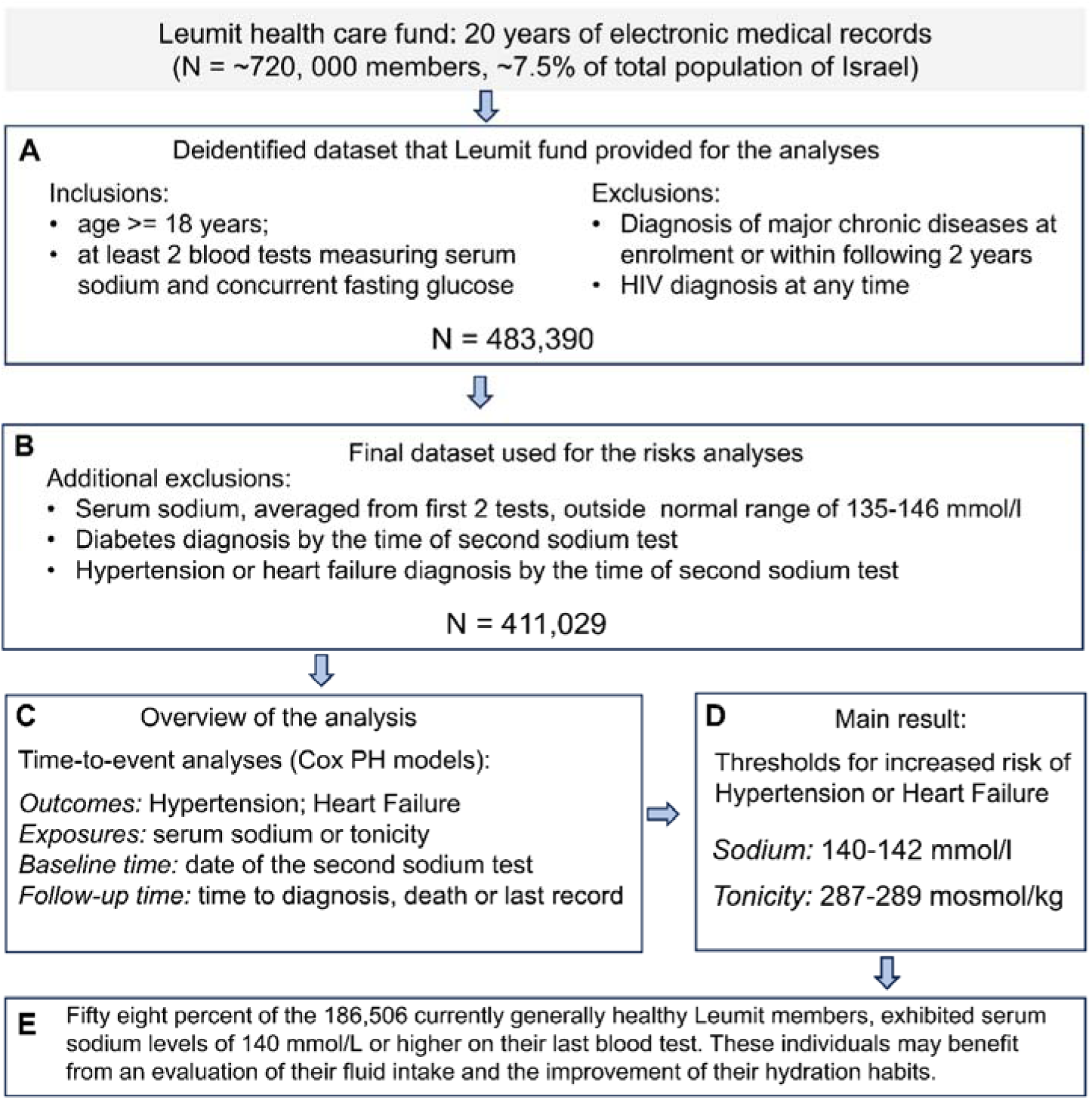
Study cohort selection, analysis, and results overview. **A)** The initial de-identified dataset from Leumit comprised 483,380 adults who were without major chronic diseases for at least 2 years post-enrollment and had a minimum of two serum sodium measurements along with a glucose test documented in their medical records. Following further exclusions **(B)**, time-to-event analyses for hypertension and heart failure were conducted, with baseline sodium levels and tonicity as the exposure variables **(C)**. The analysis identifies serum sodium and tonicity thresholds for increased risks **(D)**. The thresholds were then used to identify individuals at risk among current generally healthy Leumit members, who may benefit from evaluation of their habitual fluid intake and guidance about optimal hydration **(E)**.

Since the purpose of this analysis was to examine effects of hydration, we aimed to exclude people whose serum sodium could be affected by other factors in addition to amount of liquids they consume. For this purpose, we used the same exclusion criteria as in previous studies ^10,11^. Thus, to avoid including people with possible abnormalities of water/salt balance regulation, we excluded people who had average sodium concentration from first two tests outside normal reference range of 135–146 mmol/L and those who were diagnosed with diabetes by the time of second sodium test. To minimize reverse causation, we also excluded individuals who were already diagnosed with hypertension (ICD-9 codes 401.xxx) or HF (ICD9 - 428.x) by the time of second sodium measurement, which was used as baseline time point for the time-to-event analysis. After these exclusions, 411,029 individuals remained in the final dataset (Figure 1).

### Exposure variables

#### Serum sodium

The primary exposure variable was average serum sodium from first two blood tests available in medical records for each person. Similar to our previous studies, we performed the analysis with the assumption that the average of the two serum sodium measurements represents the hydration habits of each individual ^10,11^. Serum sodium was used as a categorical variable consisting of four groups: 135-139.5 mmol/l, 140-142 mmol/l, 142.5-143 mmol/l and 143.5-146 mmol/l. This was the same categorization that was used previously in analyses of Atherosclerosis Risk in Communities study ^10^ to see if results could be confirmed in data from general health records.

#### Serum tonicity

Our second exposure variable was serum tonicity calculated from serum sodium and glucose: (2[Na+(mmol/l)]+Glucose (mg/dL)/18).^12^ Serum sodium represents 95–98% of the serum tonicity. However, since serum tonicity is a main activator of ADH release during hypohydration leading to activation of water conservation mechanism ^13^, we included it as an additional measure of hydration status in our analysis. Serum tonicity was used as categorical variable consisting of three groups as in our previous study: 275-286 mosmol/kg, 287-289 mosmol/kg, and 289-300 mosmol/kg ^10^.

### Outcomes

Two outcomes were analyzed in the current study, incident hypertension and incident heart failure that were ascertained from first appearance of corresponding ICD-9 diagnostic codes in the medical records: 401.xxx for hypertension and 428.xxx for heart failure.

### Statistical analysis

Cox proportional hazard models were used in the time-to-event analyses to assess association of serum sodium and tonicity as measures of hydration habits with risk of future development of hypertension or heart failure. Time from baseline examination to outcome disease diagnosis was used as time variable. In absence of the outcome event, participants were censored at the time of death or last follow-up. The models were adjusted for age at second sodium test that was used as baseline, since averaged values from first and second blood tests for sodium and tonicity were used as exposure variables. The analyses were preplanned and documented in the data request submitted to Leumit prior to obtaining access to the data.

The analyses were performed using IBM SPSS Statistics version 28.0.1.1 (Armonk, NY: IBM Corp). Some plots were generated using GraphPad Prism version 9.0.2 for Windows (GraphPad Software, San Diego, California USA).

## Results

### Study population

Time-to-event analyses for hypertension and heart failure were performed on 411,129 individuals from Leumit medical records (236,295 females and 174,734 males) lacking diagnosis of major chronic diseases at baseline and who remained after exclusion of people whose serum sodium concentration could be shifted by factors affecting water balance regulation and therefore would not correctly represent level of hydration (see methods for details) (Figure 1). The characteristics of the study population are presented in Table 1. Average age at baseline, which is time of second sodium test in the medical records, was 35.09 (SD 15.68) years for females and 37.32 (15.64) years for males. The age was higher in higher sodium groups. During follow-up (up to 20 years, median (IQR): 8.54 (12.35) years for females and 12.31 (11.03) years for males), hypertension was developed by 13.3% (n=23,258) of males and 11.4% (n=26,987) of females. Heart failure developed in by 2.6%(n=4,534) of males and 1.7% (n=3,941) of females. The mean sodium levels remained almost identical between test 1 and test 2 within 4 sodium groups (Table 1) which is consistent with assumption of its stability representing individual hydration habits, as was also shown in previous studies ^14,15^. To minimize random fluctuations of test results or test day hydration, we used average serum sodium from the first two tests for analyses.

**Table 1.** Demographic and clinical characteristics of people included in the time-to-event analysis of association between baseline serum sodium and risk of hypertension and heart failure (HF)

**a) Females**
| Baseline characteristics | Females | Serum sodium groups, mmol/l (Average of 2 first tests) |  |  |  |
| --- | --- | --- | --- | --- | --- |
|  |  | 135-139.5 | 140-142 | 142.5-143 | 143.5-146 |
| no. (%) | 236,295 (100) | 106,116 (44.9) | 98,203 (41.6) | 18,113 (7.7) | 13,863 (5.9) |
| Age, years<br>– mean ( SD) | 35.09 (15.68) | 31.67 (13.55) | 35.94 (15.88) | 41.47 (17.41) | 47.01 (17.73) |
| Follow-up time, years –<br>median (IQR) | 12.3 (6.1-18.2 ) | 9.9 (5.2-17.5) | 13.3 (6.6-18.4) | 15.5 (9.8-18.7) | 16.5 (11.6-18.9) |
| Hypertension – no. (%) | 26,987 (11.4) | 8,606 (8.1%) | 11,967 (12.2%) | 3,180 (17.6%) | 3,234 (23.3%) |
| HF – no. (%) | 3941 (1.7) | 1,553 (1.3) | 2,564 (2.3) | 791 (3.8) | (863) 5.4% |
| Serum Na<sup>+</sup>, mmol/l – mean (SD) |  |  |  |  |  |
| Test 1 | 139.83 (2.42) | 138.16 (1.79) | 140.61 (1.64) | 142.39 (1.54) | 143.8 (1.72) |
| Test 2 | 139.86 (2.52) | 138.10 (1.85) | 140.67 (1.68) | 142.56 (1.57) | 144.05 (1.82) |
| Time between tests, years<br>– mean, SD | 1.96 (2.06) | 1.95 (2.12) | 2.01 (2.06) | 1.88 (1.89) | 1.72 (2.06) |

**b) Males**
| Baseline characteristics | Males | Serum sodium groups, mmol/l (Average of 2 first tests) |  |  |  |
| --- | --- | --- | --- | --- | --- |
|  |  | 135-139.5 | 140-142 | 142.5-143 | 143.5-146 |
| no. (%) | 174,734 (100) | 49,953 (28.6) | 86,185 (49.3) | 21,153 (12.1) | 17,443 (10.0) |
| Age, years<br>– mean ( SD) | 37.32 (15.64) | 37.13 (15.97) | 36.52 (15.29) | 38.33 (15.49) | 40.76 (16.05) |
| Follow-up time, years –<br>median (IQR) | 12.3 (6.8-17.8) | 10.3 (6.1-17.8) | 12.2 (6.5-17.8) | 14.0 (8.4-17.9) | 14.8 (9.8-18.0) |
| Hypertension – no. (%) | 23,258 (13.3%) | 6,000 (12.0%) | 11,021 (12.8%) | 3,096 (14.6%) | 3,141 (18.0%) |
| HF – no. (%) | 4,534 (2.6%) | 1,863 (3.2%) | 2,866 (2.8%) | 833 (3.5%) | 825 (4.2%) |
| Serum Na<sup>+</sup>, mmol/l – mean (SD) |  |  |  |  |  |
| Test 1 | 140.62 (2.40) | 138.39 (1.75) | 140.78 (1.62) | 142.51 (1.56) | 143.94 (1.76) |
| Test 2 | 140.56 (2.42) | 138.37 (1.81) | 140.68 (1.63) | 142.46 (1.59) | 143.99 (1.86) |
| Time between tests, years<br>– mean, SD | 2.47 (2.52) | 2.41 (2.58) | 2.57 (2.57) | 2.46 (2.44) | 2.21 (2.19) |

### Serum sodium and tonicity and risk of hypertension in Leumit medical records cohort

To assess the association of serum sodium and tonicity with risk of hypertension in the Leumit medical records cohort, we performed time-to-event analysis using Cox proportional hazard models (Figure 2, Supplemental Tables 1, 2). After adjusting for age and sex, the risk to develop hypertension gradually increased with increasing sodium concentration. The risk was already elevated by 12% in the 140-142 mmol/l group (HR 1.12, P<0.0001) relative to 135 -139.5 mmol/l reference group and reached 30% in 143.5-146 mmol/l group (HR 1.30, P<0.0001). Similarly, the risk to develop hypertension increased with rising tonicity reaching 19% increased risk in 289-300 mosmol/kg group (HR 1.19, P<0.0001) relative to 275-287 mosmol/kg reference group (Figure 2B, 2D, Supplemental Table 2). The risks were similar for both females and males, increasing up to 27% (HR 1.27, P<0.0001) for 143.5 -146 mmol/l sodium group (Figure 2A, 2C, Supplemental Table 1) and up to 17-19% (P<0.0001) in 289-300 mosmol/kg group. In models adjusted for age and sex, for both sodium and tonicity, males showed about 7% higher risk to develop hypertension (Figure 2, Supplemental Tables 1, 2).

**Figure 2.**
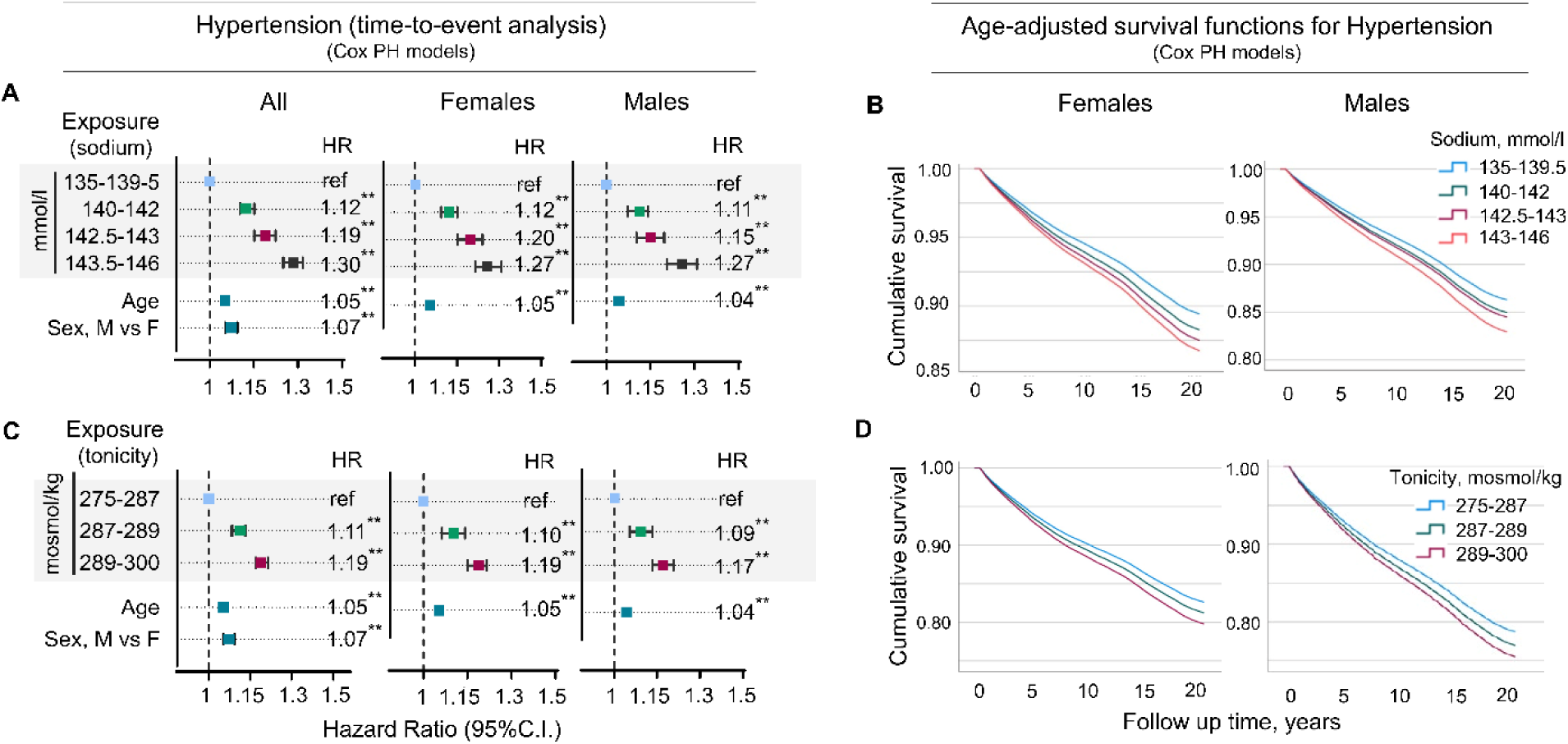
Serum sodium and tonicity at baseline and risk of hypertension among Leumit members. **A,B)** Assessment of relative risk to develop hypertension in four sodium groups. **A)** Time-to-event analysis. Cox proportional hazard models run for whole cohort and separately for females and males. *Individuals with serum sodium within 135-139.5 mmol/l sodium group have lowest risk to develop hypertension. Risk increases at sodium levels of 140 mmol/l and higher. See also Supplemental table 1for full Cox-PH model results;* **B)** Cumulative survival curves for hypertension for females and males. **C,D)** Assessment of relative risk to develop hypertension in three tonicity groups. **C)** Time-to-event analysis. Cox proportional hazard models run for whole cohort and separately for females and males. *Individuals with serum tonicity within 275-287 mosmol/kg tonicity range have lowest risk to develop hypertension. Risk increases at tonicity levels exceeding 287 mosmol/kg. See also Supplemental table 2 for full Cox-PH model results;* **D)** Cumulative survival curves for hypertension for females and males.

### Serum sodium and tonicity and risk of heart failure in Leumit medical records cohort

To assess association of serum sodium and tonicity with risk of heart failure in the Leumit medical records cohort, we performed time-to-event analysis using Cox proportional hazard models (Figure 3, Supplemental Tables 3, 4). After adjusting for age and sex, the risk of developing HF was already increased in 140-142 mmol/l sodium group and 287-289 mosmol/kg tonicity group. It reached a 20% increase in 143.5-146 mmol/l sodium group (Figure 3A, C, Supplemental Table 3) and 16% increase in 289-300 mosmol/l tonicity group (Figure 3B, D, Supplemental Table 4) compared to the reference groups of 135-139.5mmol/l for sodium and 275-287 mosmol/kg for tonicity. While the sodium/tonicity dependence for hypertension was similar for females and males (Figure 2, Supplemental Tables 1, 2), there was substantial sex difference in the risk of HF. For females, the risk of developing HF was already increased in 140-142 mmol/l sodium group reaching 28% (P<0.0001) in 143.5-146 mmol/l sodium group. For males, there was a trend for increased risk up to 9% in 143.4-146 mmol/l sodium group, but it reached significance only in models with serum tonicity as exposure variable (Figure 3, Supplemental Tables 3, 4). In models adjusted for age and sex, for both sodium and tonicity, males showed about 50% higher risk of developing heart failure (Figure 2, Supplemental Tables 3, 4).

**Figure 3.**
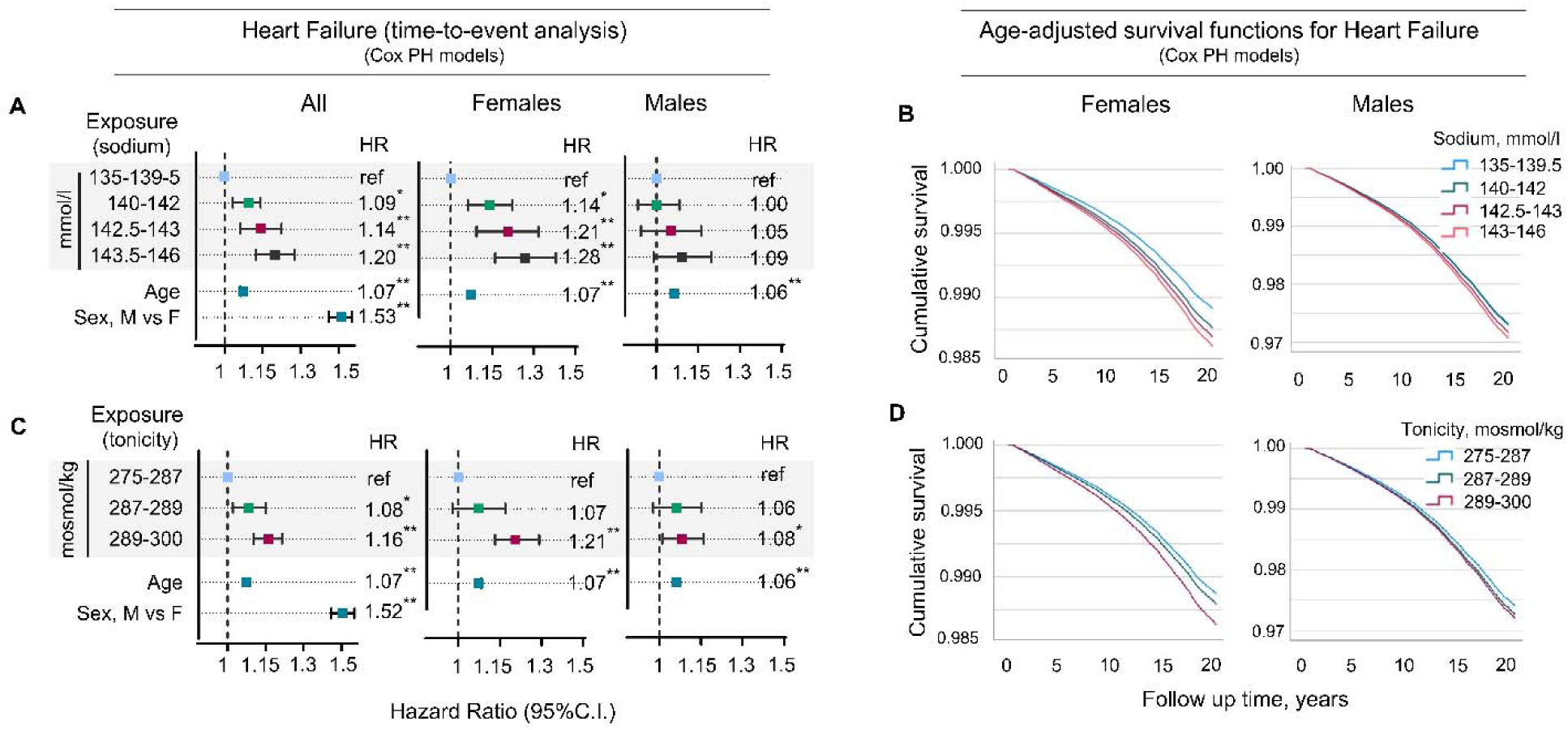
Serum sodium and tonicity at baseline and risk of heart failure among Leumit members. **A,B)** Assessment of relative risk to develop heart failure in four sodium groups. **A)** Time-to-event analysis. Cox proportional hazard models run for whole cohort and separately for females and males. *Individuals with serum sodium within 135-139.5 mmol/l sodium group have lowest risk to develop heart failure. Risk increases at sodium levels of 140 mmol/l and higher for females but not for males. See also Supplemental table 3 for full Cox-PH model results;* **B)** Cumulative survival curves for heart failure for females and males. **C,D)** Assessment of relative risk to develop heart failure in three tonicity groups. **C)** Time-to-event analysis. Cox proportional hazard models run for whole cohort and separately for females and males. *Risk to develop heart failure increases when serum tonicity exceeds 287 mosmol/kg for females, and 289 mosmol/kg for males. See also Supplemental table 4 for full Cox-PH model results;* **D)** Cumulative survival curves for heart failure for females and males.

### Using serum sodium for identification of people at risk among current Leumit members

In the analysis presented in previous sections, we analyzed 20 years of medical records from both past and present Leumit Health Services members. Through this analysis, we identified serum sodium levels that were associated with elevated risk of developing hypertension or heart failure within this cohort. We observed a slight increase in risk (up to 12%) for sodium levels ranging from 140-142 mmol/L, which escalated to a more significant risk (up to 30%) at levels between 142-146 mmol/L (Figures 2 and 3). These sodium thresholds may be used in clinical practice for identifying individuals at increased risk.

In subsequent analyses, we estimated the number of current Leumit members at increased risk based on their most recent sodium test results, who might benefit from adopting better hydration practices. We applied the same inclusion and exclusion criteria as in our earlier risk analysis. Therefore, we included only those currently enrolled members who were generally healthy, which we defined as not having a diagnosis of major chronic diseases, including diabetes, hypertension and heart failure, at the time of their last sodium test and having serum sodium levels within the normal range (135-146 mmol/L) (Figure 4A).

**Figure 4.**
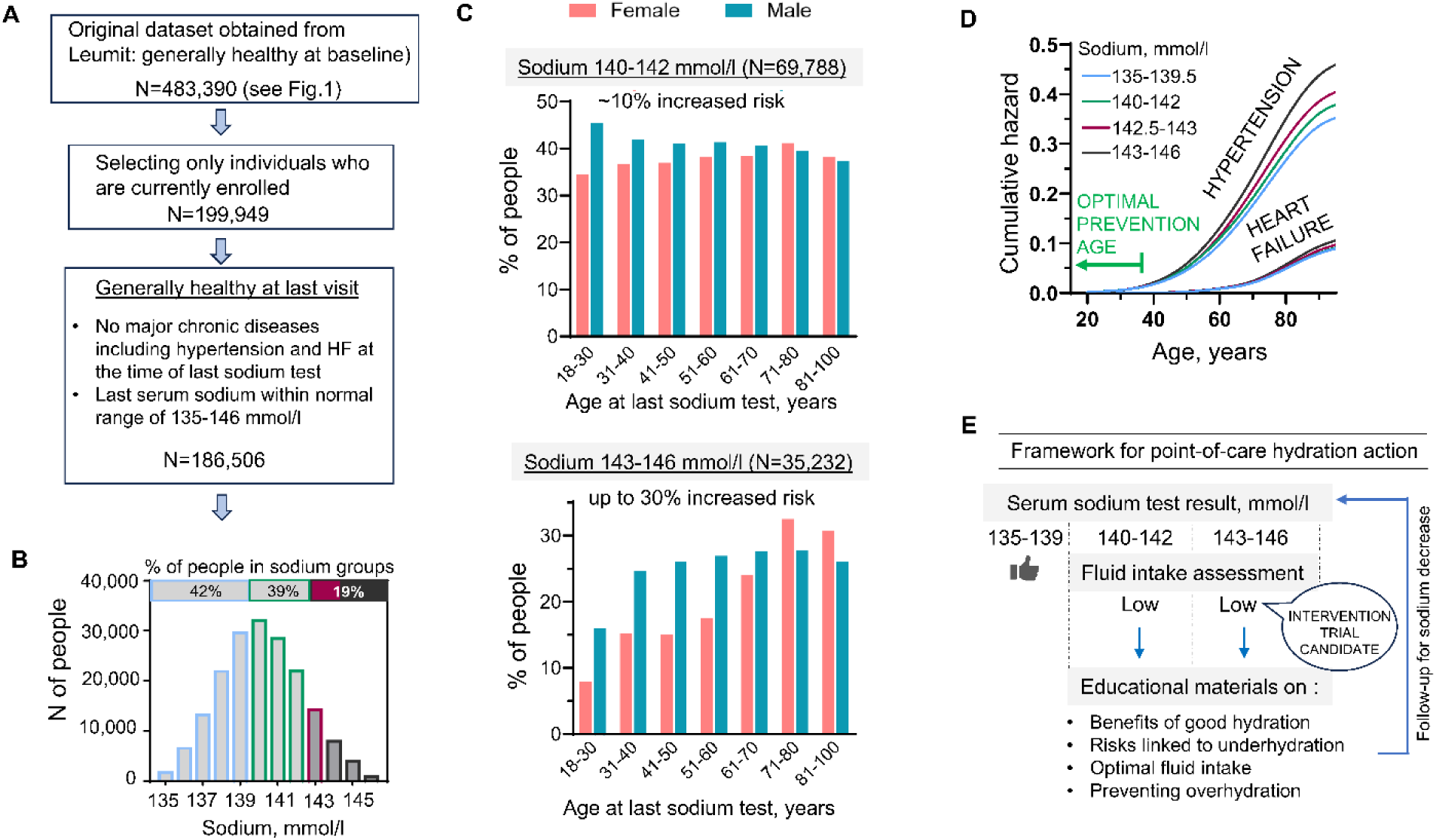
Identification of at-risk individuals based on serum sodium levels in current Leumit health fund members, who may benefit from hydration-focused intervention. **A)** Selection of generally healthy individuals lacking diagnosis of major chronic diseases including hypertension and heart failure (N=186,506). **B)** Distribution of serum sodium levels among selected individuals shows that 58% fall within the 140-146 mmol/L range, which is associated with an elevated risk. **C)** Proportion of individuals with serum sodium levels in the ranges of 140-142 mmol/l (upper panel) and 143-146 mmol/l (lower panel), stratified by age and sex. **D)** Cox cumulative hazard curves for onset of hypertension and heart failure by sodium groups. Age from first diagnosis serves as the time variable. Plots from the Figure 1B cohort show risk accumulation, guiding optimal timing for early interventions, ideally before age 40. **E)** Framework for point-of-care hydration interventions. A serum sodium level of 140 mmol/L or above may indicate the need to assess habitual fluid consumption. If intake is below recommended levels, patients receive advice on optimal hydration and educational materials to avoid overhydration. The effectiveness of these interventions is monitored through subsequent serum sodium tests.

Out of 186,506 individuals who met these criteria, 72,897 (39.1%) had their last sodium test results in the 140-142 mmol/L range, which is associated with a slightly increased risk of developing hypertension and heart failure. Moreover, 36,167 (19.4%) had results in the 143-146 mmol/L range, which carries a substantially higher risk of up to 20-30% (Figures 4A,B). Depending on age and sex, the percentage of individuals with sodium levels in the 143-146 mmol/L group varied from 7.9% to 32.5% (Figure 4C). Additionally, depending on age and sex, another 34.6 % to 45.5% of individuals had sodium levels in the 140-142 mmol/L range (Figure 4C), linked to up to 12% increased risk for hypertension and heart failure (Figures 2, 3). The estimation of the age-dependent probability of developing hypertension and heart failure among Leumit members indicated that the onset of hypertension began to rise in the late thirties, while heart failure began to increase in the late fifties, as shown in Figure 4D. This suggests that while preventive interventions are beneficial at any age, they would likely be most effective if implemented before the age of 40.

In summary, among the current generally healthy members of Leumit Health Services, an estimated 53% of females and 66% of males have serum sodium levels within the 140-146 mmol/L range. This group could benefit from evaluation of their hydration habits and guidance regarding adequate fluid intake to maintain optimal hydration (Figure 4E).

## Discussion

Electronic medical records have become commonplace in numerous countries worldwide. As we accumulate data with adequate follow-up times, we can identify region- and culture-specific health trends and associations. This information will provide healthcare professionals with community-specific insights and inform public health interventions aimed at improving health outcomes in these areas.

In the current study, we analyzed data from historical and ongoing clinical records collected in Israel by Leumit Healthcare Services over a 20-year period, encompassing 700,000 past and present members.

The analysis had two objectives. The first was to seek further validation for the association between high-normal serum sodium levels and the risk of chronic diseases, as identified in epidemiological studies ^10,11,16,17^, by using a different cohort and utilizing data collected from routine healthcare practices. The second objective was to explore how electronic records generated within regular healthcare systems can facilitate identification of at risk individuals who might benefit from implementation of hydration-focused interventions.

### Verifying link between serum sodium and chronic diseases in Leumit electronic medical records

We assessed the risk of hypertension and heart failure in relation to serum sodium levels within the normal range of 135-146 mmol/L. Categorization of sodium was based on previous analyses of 15,000 middle-age (45-65 years) individuals from the ARIC study—a US population-based cohort, that demonstrated an increased risk of heart failure, other chronic diseases and premature mortality among participants with high-normal serum sodium levels ^10,11^.

The trend toward a heightened risk of heart failure at high-normal serum sodium levels was consistent with findings from the ARIC study. However, current analysis identified lower threshold for the increased risks. The risks already became apparent in the 140-142 mmol/L sodium group (Figure 3), while in the ARIC cohort, they started at 142 mmol/L ^10^. The difference between these thresholds may be due to the increased statistical power which results from a larger sample size (400,000 individuals compared to 15,000 in the ARIC cohort), the wider age range (18-90 years, as opposed to 45-65 years in the ARIC study), or other potential factors including variations in ethnicity, and local environmental and climatic conditions. In the present study, we not only confirmed the association of sodium level group with heart failure (Figure 3) but also delineated an association with risk to develop hypertension (Figure 2). Since hypertension is the most common risk factor for chronic diseases and is the leading contributor to all-cause mortality and disability worldwide ^5,18^, reducing its risk through hydration-focused interventions could provide further preventive benefits ^4^.

The risk for both hypertension and heart failure was already slightly increased (up to 12%) at sodium levels of 140-142 mmol/l and tonicity of 287-289 mosmol/kg and becomes more pronounced (up to 30%) when these levels exceed 142 mmol/L for sodium and 289 mosmol/kg for tonicity (Figures 2 and 3). These thresholds align precisely with those that trigger the secretion of arginine vasopressin (AVP), also known as antidiuretic hormone (ADH), from the pituitary gland, which then activates the reabsorption of water by the renal collecting duct, leading to the excretion of a reduced volume of more concentrated urine. ^19^. The heightened risk of hypertension and heart failure at levels of body water content that triggers AVP secretion may be due to the chronic activation of the integral physiological response to decreased body water content aimed at conserving water and adjusting the volume of the vascular system through vasoconstriction to maintain blood pressure and tissue perfusion (reviewed in ^6^). This result suggests that optimal hydration, which is associated with minimal risks of hypertension and heart failure, represents a state of water balance where serum sodium and osmolality are maintained below the threshold that triggers AVP-stimulated water conservation mechanisms.

### Considerations for designing hydration-focused intervention

Currently, no established protocols or guidelines exist that outline how to implement long-term hydration interventions at the population level with the aim of preventing the development of chronic diseases by maintaining optimal hydration. However, several interventional clinical trials have assessed the effect of improved hydration on the progression of established diseases, including chronic kidney disease, polycystic kidney disease, recurrent urinary tract infections, and kidney stone formation ^20-22^. Additional trials have begun to focus on hydration in healthy individuals ^8,23^. Challenges encountered by these trials provide valuable information for designing future hydration-focused interventions (reviewed in ^6^).

A key challenge that the interventional trials faced is ensuring that participants consistently drink more water. The identified barriers to increased water intake included lack of awareness of the potential benefits, forgetting to drink, disliking the taste of water, absence of thirst, limited access to water, and increased urination disrupting work^24^. For instance, despite three reminder calls in a study where participants were to drink an additional 1.5 liters daily for six months, the increase in water consumption was a mere 350ml per day compared to the control group ^25^. Another study, despite more diligent tracking and coaching, saw only a 0.6-liter increase in daily urine output ^22^. To overcome these issues, we need innovative, interactive daily strategies that highlight the benefits and motivate appropriate water intake.

Another key lesson is the importance of targeted participant selection to observe the benefits of hydration interventions. Effective studies should focus on individuals who typically consume little water and exhibit markers indicating low hydration levels (Figure 4E). The beneficial effects of hydration are most apparent when such specific criteria guide participant selection ^26^.

When considering hydration interventions, safety issues always come into play. Advising people to drink more fluids is linked to concerns about the dangers of fluid overconsumption, which can lead to dilutional hyponatremia and can be fatal ^27^. Concerns about dangers of fatal water intoxication for healthy individuals are often overestimated and can be addressed through educational information. The maximal excretion rate of even minimally diseased kidneys is 0.8–1 L per hour, meaning that kidneys can excrete up to 24 L of water load a day that by far exceeds general recommendations ^27-31^. To prevent water intoxication, the rate of water gain should not exceed the maximal excretion rate. Most cases of hyponatremia caused by excessive drinking occur when a large amount of water is consumed over a short period of time, exceeding one liter per hour ^31,32^. Providing this information alongside with recommendations for optimal fluid intake will help to avoid overhydration (Figure 4E). All of the studies performed so far have concluded that a prescribed increase of 1–3 l of fluid intake per day on top of habitual intake is safe in healthy individuals. Moreover, no adverse effects of increased water consumption occurred even in individuals with established kidney diseases who participated in these trials^21,22,25^.

Additional challenge in implementing hydration interventions is the expected variations in fluid intake requirements that may be needed to achieve optimal hydration across different global locations, influenced by environmental factors such as latitude, altitude, air temperature, and humidity, as well as local traditions and lifestyles. A recent study that analyzed water turnover in individuals from 23 countries using isotope-tracking methods highlighted these regional and lifestyle differences ^33^. This suggests that tailored adjustments to both risk assessment criteria and fluid intake recommendations may be necessary to effectively promote optimal hydration in diverse populations worldwide.

### Identifying at-risk individuals among current Leumit members

The data on risk thresholds obtained from the 20-years of Leumit electronic health records (as shown in Figures 1, 2, 3, and 4D) are directly relevant to the current members of the health plan, representing a subset of the original cohort used for the analysis and target population for hydration intervention (Figure 4). Our analysis of current 186,506 Leumit members who are presently free from diagnosis of major chronic diseases including hypertension and heart failure showed that a significant portion of them - 39% or 72,897 individuals - have serum sodium levels between 140-142 mmol/L, placing them at up to a 12% increased risk for hypertension and heart failure. Furthermore, 19% of members, which equates to 36,167 individuals, fall into a higher risk category with sodium levels between 143-146 mmol/L, with up to 30% increased risk. These numbers indicate that a large proportion of the population might substantially reduce their health risks through better hydration practices, underscoring the widespread potential benefits of improved hydration (Figure 4E).

Maintaining optimal hydration is a lifelong endeavor. Utilizing electronic medical records within general healthcare provides a means to address many of the challenges faced in clinical trials. By using these records, healthcare providers can easily identify suitable candidates for hydration intervention and track the impact of hydration on health without the burden of collecting extensive medical histories or scheduling extra appointments. Educating patients on achieving proper hydration can be efficiently accomplished within general healthcare systems, where physicians maintain long-term relationships with patients. Serum sodium testing is routinely performed and widely accessible. If further research confirms that hydration optimization can lower serum sodium levels, this metric may serve as a reliable indicator of successful hydration interventions.

## Acknowledgments

The authors would like to thank the staff of Leumit Health Services and Leumit Start for creating and maintaining electronic medical records, and providing access to the data for our analysis.

## Data availability statement

Electronic medical records data provided for analysis by the Leumit Health Services cannot be shared with third parties.

## Contributions

JR initiated the study. JR and NID conceptualized the study, wrote and edited the original draft. JR conducted the statistical analysis in collaboration with NID. NID also prepared figures and tables. JR was also involved in funding acquisition and project administration. MD contributed to conceptualizing the study, validating the data and project administration. NB curated and validated the data, managed resources, advised on aspects of methodology relating to understanding the data. MB contributed to study conceptualization, funding acquisition and reviewing and editing. NB and JR had direct access to the data. JR and NID contributed equally.

## Structured Graphical Abstract

An analysis of 20 years of electronic medical records from a general healthcare plan showed that healthy individuals with high normal serum sodium exceeding 140 mmol/l and tonicity exceeding 287 mosmol/kg have an increased risk of developing hypertension and heart failure. This risk may be induced by a physiological response to underhydration, activated by the elevated sodium and tonicity levels. The analysis estimated that 58% of currently healthy members may benefit from optimal fluid intake guidance based on their most recent serum sodium test result. Worldwide surveys found that over 50% of people do not meet daily fluid intake recommendations, suggesting that preventive, hydration-focused interventions in primary healthcare could have a significant impact. ADH – antidiuretic hormone; RAAS - Renin-Angiotensin-Aldosterone System; SNS – Sympathetic Nervous System; EMR – electronic medical records. Created with BioRender.com.

### Key Question

Can serum sodium levels in general care electronic medical records identify generally healthy individuals at increased risk of hypertension and heart failure during routine physical exams, who might benefit from guidance on optimal fluid intake?

### Key Finding

An analysis of 20 years of electronic medical records from 411,029 adults registered with Israel’s Leumit Healthcare Services, who were healthy at baseline, showed an increased risk of hypertension and heart failure in those with serum sodium levels between 140-146 mmol/L compared to 135-138 mmol/L.

### Take-home Message

Electronic medical records could be utilized to identify individuals at increased risk of hypertension and heart failure based on serum sodium levels, suggesting that preventive interventions focused on hydration could be implemented within primary healthcare settings.

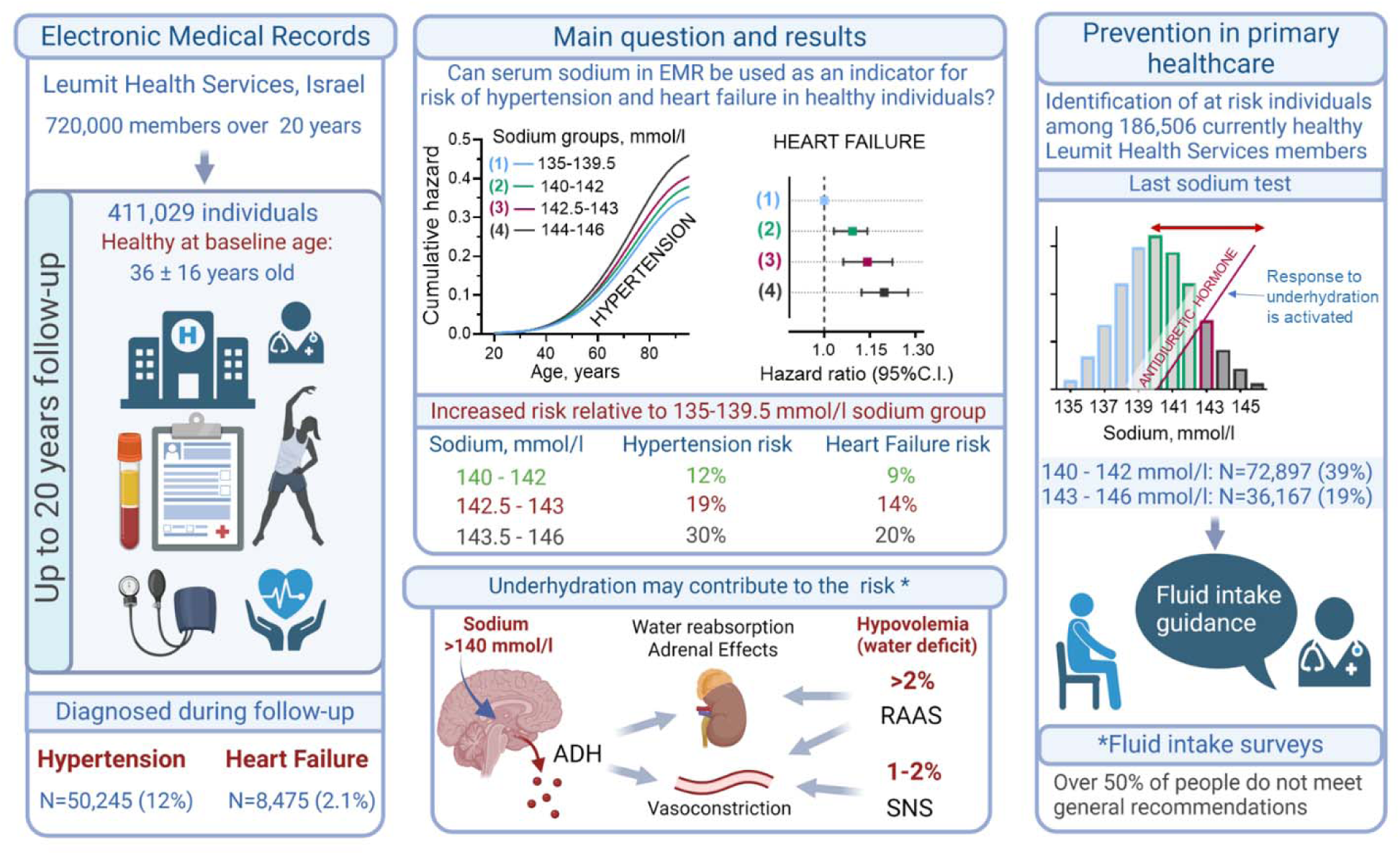

